# Enhancing Semantic Segmentation in Chest X-Ray Images through Image Preprocessing: ps-KDE for Pixel-wise Substitution by Kernel Density Estimation

**DOI:** 10.1101/2024.02.15.24302871

**Authors:** Yuanchen Wang, Yujie Guo, Ziqi Wang, Linzi Yu, Yujie Yan, Zifan Gu

## Abstract

**Background:** Deep-learning-based semantic segmentation algorithms, in combination with image preprocessing techniques, can reduce the need for human annotation and advance disease classification. Among established preprocessing techniques, CLAHE has demonstrated efficacy in enhancing the segmentations algorithms across various modalities.

**Method:** This study proposes a novel preprocessing technique, ps-KDE, to investigate its impact on deep learning algorithms to segment major organs in posterior-anterior chest X-rays. Ps-KDE augments image contrast by substituting pixel values based on their normalized frequency across all images. Our approach employs a U-Net architecture with ResNet34 (pre-trained on ImageNet) serving as the decoder. Five separate models are trained to segment the heart, left lung, right lung, left clavicle, and right clavicle.

**Results:** The model trained to segment the left lung using ps-KDE achieved a Dice score of 0.780 (SD=0.13), while that trained on CLAHE achieved a Dice score of 0.717 (SD=0.19), *p*<0.01. ps-KDE also appears to be more robust as CLAHE-based models misclassified right lungs in select test images for the left lung model.

**Discussion:** Our results suggest that ps-KDE offers advantages over current preprocessing techniques when segmenting certain lung regions. This could be beneficial in subsequent analysis such as disease classification and risk stratification.

## 1. Introduction

With recent advances in artificial intelligence, deep-learning (DL) has emerged as a leading machine learning technique in medical imaging analysis, playing a transformative role in tasks such as image segmentation(1, 2, 3). This capability extends to various applications, including the segmentation of breast lesions(4, 5), classification of pulmonary cancer stages(6), tissue characterization(7), detection of cardiomegaly(8), and many more. The improved performance for these intricate tasks suggests the potential of computer-aided techniques to improve diagnosis via segmentation.

Within the context of radiology, the segmentation of organs and tumors in medical images holds promise for disease diagnosis and treatment(7). Despite the improved performance in intricate tasks, the effectiveness of deep learning, with its efficient feature representation learning, is contingent upon extensive training data(7). This dependency, however, has spurred the evolution of novel image segmentation approaches. One such milestone was the fully convolutional network (FCN), pioneering pixel-to-pixel semantic segmentation(9). FCN’s innovation lies in replacing the last fully connected layer with a deconvolutional layer. Building upon this foundation, the U-Net model as a modification of FCN increases the number of deconvolutional layers and therefore effectively captures more context while requiring smaller training smaples(10). Notably, U-Net has found wide-spread application in segmenting medical images across various modalities, including X-rays, MRI, CT, and pathological images(3, 11, 12).

Image enhancement, prior to the introduction of DL algorithms, played a crucial role in studying medical images by providing human viewers with crucial information to facilitate analysis(13). Among the techniques employed, adaptive histogram equalization (AHE) emerged as a popular image preprocessing method that splits each image into tiles where the histogram of each tile is remapped to smooth tile boundaries(13, 14, 15). An enhancement upon traditional AHE methods, contrast limited adaptive histogram equalization (CLAHE) was introduced by clipping histograms to constrain the contrast(16). Recently, several studies have shown the advantages of CLAHE on DL-related tasks, such as predicting five stages of diabetic retinopathy(17), segmenting temporomandibular joint articular disks from magnetic resonance images(18), and classification of COVID-19 and other pneumonia cases(19).

In this study, we propose a novel, histogram-based, image preprocessing method termed ps-KDE, aiming to assess its impact on segmentation algorithms applied to the anatomic structure of chest X-ray images. ps-KDE brings three notable contributions: 1) it presents an end-to-end data augmentation method characterized by its simplicity of implementation and adaptability for fine-tuning to accommodate diverse datasets; 2) it demonstrates the efficacy of a density-based augmentation method in segmenting vital organs in chest X-rays; and 3) it establishes the robustness of segmentation algorithms through the interpretation of heatmaps generated by the model.

## 2. Data and Methods

### 2.1. Data

We used a publicly available database with 247 posterior-anterior (PA) chest radiographs collected from 13 institutions in Japan and one in the United States. The original radiographs are provided by the Japanese Society of Radiological Technology (JSRT) Database(20) and the manual mask annotations are provided by the Segmentation in Chest Radiology (SCR) Database(21). The chest radiographs are in PNG format, and the labels are in the form of binary masks. Each image in the database was scanned from film to a size of 2048*2048. Among the 247 images, 154 of them showed solitary pulmonary lung nodule, while the remaining 93 images exhibited no signs of lung nodules. The ethnic representation is unknown.

Among the subset of patients with nodules, gender distribution was observed as 68 males and 86 females. In contrast, among patients without nodules, the gender distribution consisted of 51 males and 42 females. The mean age for patients with nodules is 60 years old. Each image has five matching masks generated manually by expert radiologists. Each binary mask delineates the boundary of one of the five anatomical structures: heart, left lung, right lung, left clavicle, and right clavicle. Since the original images are in grayscale, we added a singleton dimension to them, namely a single-color channel. This is necessary because our deep learning model expects the input to have color channels.

This study utilizes exclusively publicly available data and thus do not require Institutional Review Board (IRB) review per regulations set by the Office for Human Research Protections (OHRP) within the U.S. Department of Health and Human Services. The data was accessed on the third day of April of 2022. The authors had no access to information that could identify individual participants during or after data collection.

### 2.2. Data Augmentation

Large quantities of data are often needed to train most deep learning algorithms successfully. Data augmentation is crucial when large datasets are not feasible in order to prevent overfitting and increase model performance. Five types of augmentation were simultaneously applied to each of the original images and its corresponding mask so that the masks correctly represent the anatomical structures on the augmented images. Augmentations include rotation, horizontal flip, vertical flip, a range for image zooms, and rescale. The rotation can occur between 90 degrees clockwise and counterclockwise of the original orientation. Horizontal and vertical flips occur at a probability of 0.5. The range of zoom is between 0.5 and 1.5 for the original images. All images are then rescaled from the red-green-blue scale [0, 255] to [0,1] and resized to 256×256 pixels to help the predictive models achieve faster convergence and higher stability. **Figure 1** contains examples of augmentation. Data augmentation was implemented with *ImageDataGenerator* from *TensorFlow*(*22*).

**Figure 1.**
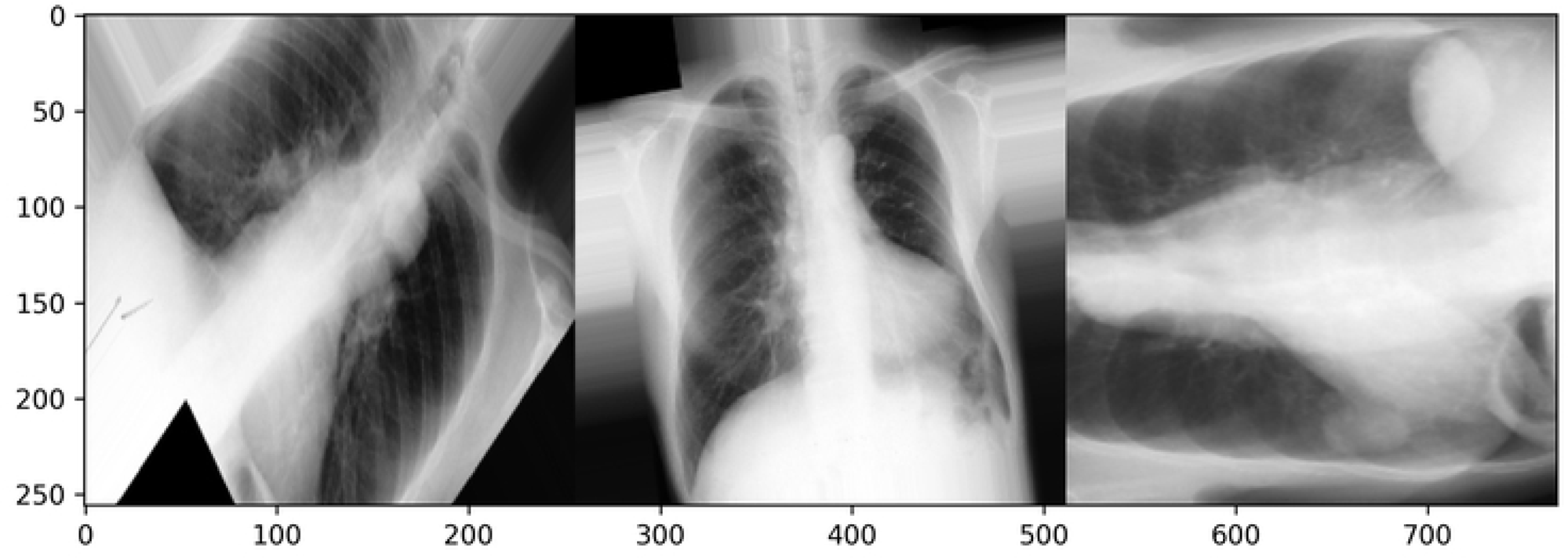
Augmented images of three distinct individuals in the training set.

### 2.3. Image preprocessing

### 2.3.1. Contrast Limited Adaptive Histogram Equalization (CLAHE)

Histogram equalization (HE) is a widely used digital image processing method to enhance the contrast of images. It expands an image’s distribution range, as some images might only occupy a small portion of the entire value range. The resulting distribution of the pixel value would become more similar to a uniform distribution. However, as most images usually use the whole range of intensity (for instance, 0-255 for a standard RGB image), the HE method would not have much impact on the image. The adaptive histogram equalization (AHE) method, as a result, was developed to overcome the short come(23). In AHE, images are divided into subsections, and each subsection is equalized separately. Compared to regular HE, AHE enhances local contrast but with the risk of over-amplifying noise in some regions. One of the improved versions of AHE, the CLAHE, clips the outliers in histograms and redistributes the values across the value range(16). An example of chest X-rays preprocessing with CLAHE is shown in **Figures 2a, 2b**. The equalization of histograms can be visualized in **Figures 3a, 3b**. The distribution of pixel values became more uniform after CLAHE.

**Figure 2.**
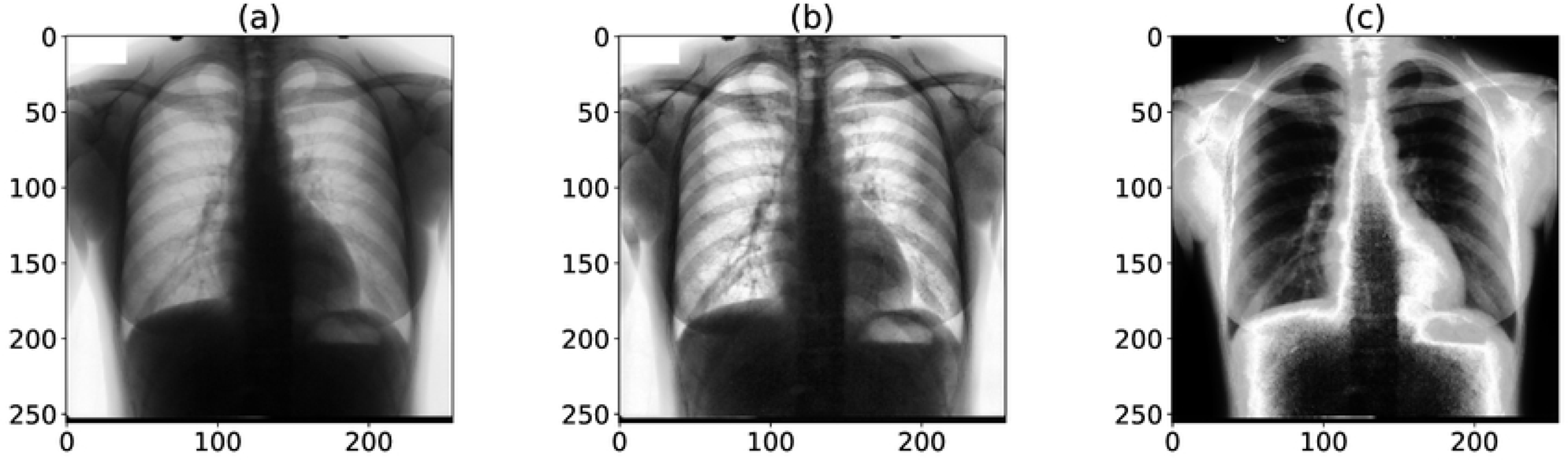
An example of an X-ray image being processed. (a) Original chest X-ray image. (b) CLAHE processed chest X-ray image. (c) ps-KDE processed chest X-ray image (location: heart).

**Figure 3.**
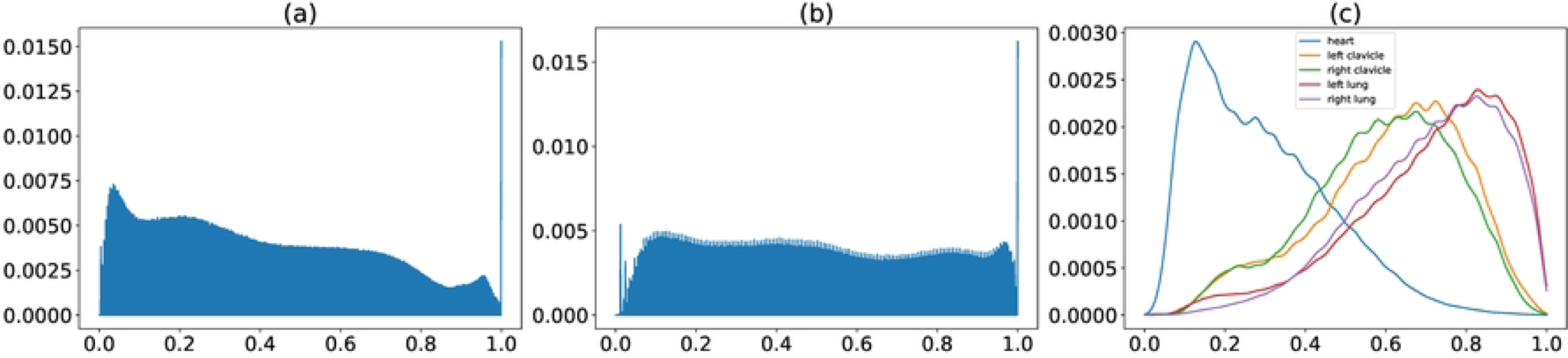
Normalized distribution of pixel values in X-ray images. (a) Histogram of original images. (b) Histogram of CLAHE-processed images. (c) KDE of pixel values in different organs.

### 2.3.2. Pixel-wise substitution by Kernel Density Estimation (ps-KDE)

During the initial exploration of the data, we observed that the distribution of pixel values appeared to be different from organ to organ. We generated histograms of pixel values in different organs to validate our initial observation. We then performed kernel density estimation (KDE) to get a probability density function (PDF) for each organ (*Algorithm 1*, **Figure 3c**). The PDFs were calculated based on the training set and were stored as prior knowledge. For each image, we substitute each pixel with the density of that pixel value (*Algorithm 2*). The image would then be mapped to a 0-1 range to ensure consistency among images. In other words, our proposed ps-KDE substitute pixel value for frequency, so that more frequently occurring pixel values in an organ would have a higher value in the resulting plot. Similar to CLAHE, the results were visually appealing (**Figure 2c**).

#### Algorithm 1: Algorithm for generating estimated probability density function

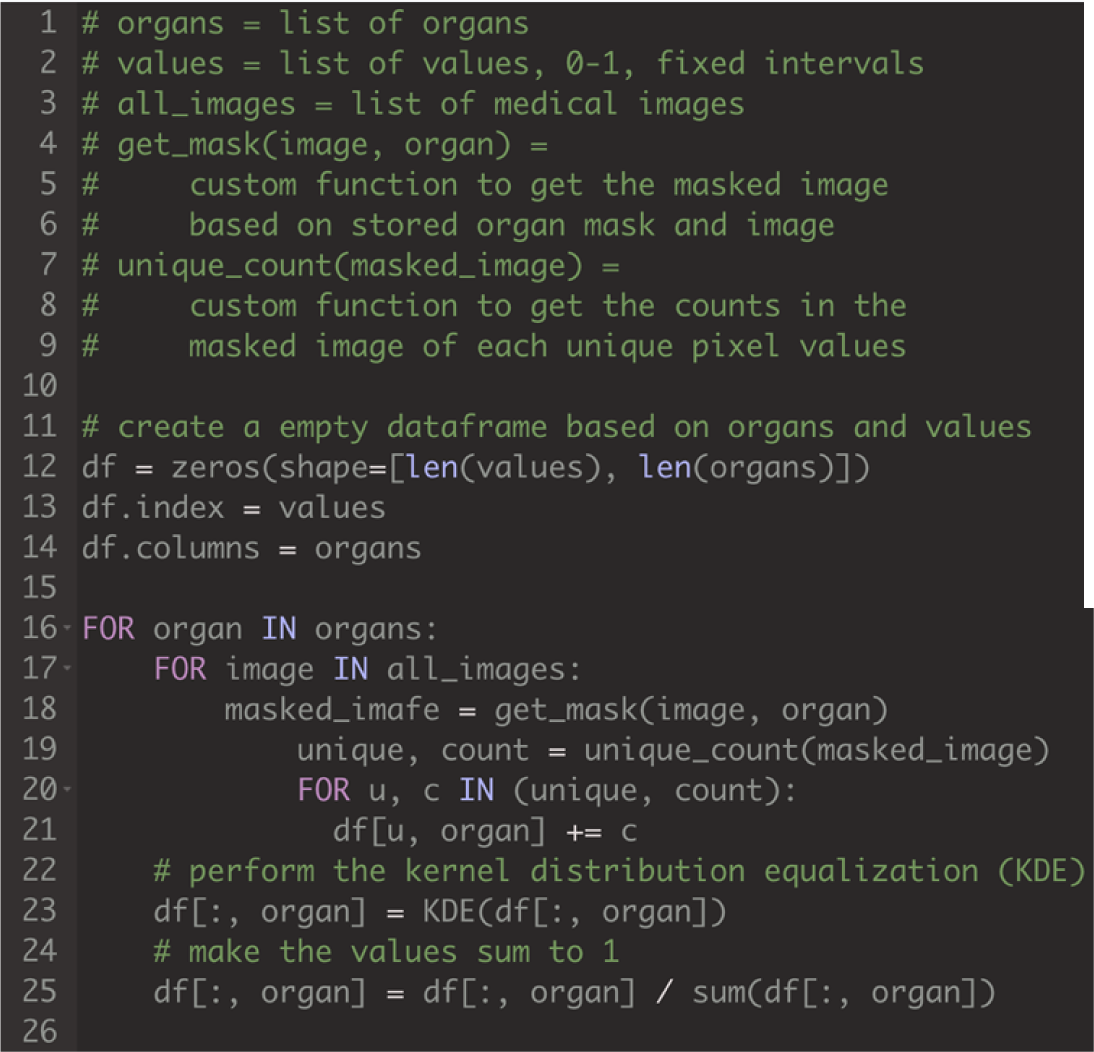

#### Algorithm 2: Algorithm for substituting pixel value with density

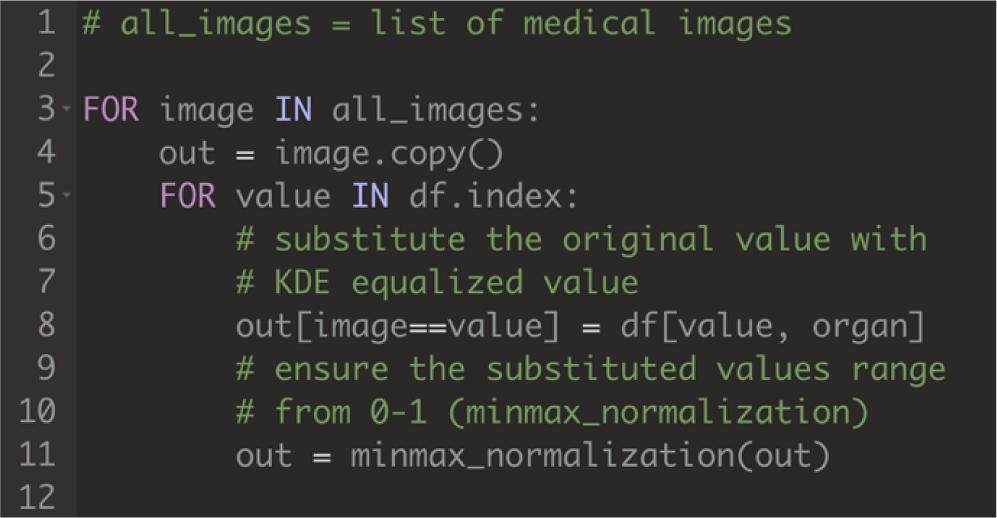

### 2.4. Model development

We employed a deep learning method for the semantic segmentation of chest radiographs, leveraging the *U-Net* neural network designed for segmentation tasks(10). The network architecture consists of a contracting path and an expansive path. The original design for the contracting path consists of unpadded convolutions with size 3×3, followed by rectified linear units with a 2×pooling layer, whereas the expansive path applies upsampling for each feature map from the contracting path to restore the original input size. The final layer maps the feature vector to the number of classes.

Implementation-wise, we used the Python package *segmentation_models* (Yakubovskiy, 2019, v1.0.1) with a *ResNet34* backbone featuring pre-trained weights from *ImageNet* (**Figure 4**). *ResNet* won the *ImageNet* Large Scale Visual Recognition Challenge 2015 with a top-five test error of 3.567 percent in the image classification category(24). With a network depth of 152, *ResNet* surpasses *VGGNet* in depth by eight times(25). Referred to as the *ResNetUnet* model in our paper, this amalgamation of *U-Net* and *ResNet34* structures incorporates additional enhancements such as batch normalization and zero padding to complement the original design.

**Figure 4.**
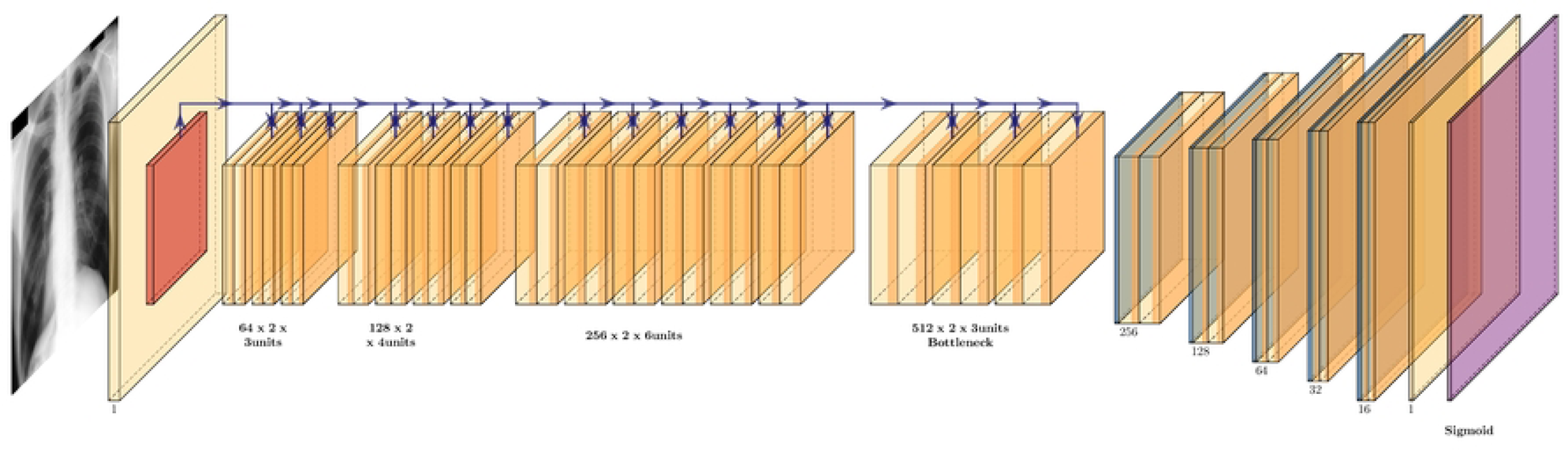
*ResNetUnet* model. *U-Net* Model Architecture Implemented with *ResNet34* Backbone

A loss function is needed for machine learning models to learn through propagation. For image segmentation tasks, multiple different loss functions could be used. For example, three loss functions were proposed to have good performances: binary cross entropy (BCE), binary cross entropy with Jaccard loss (BCE+JCD), and Dice loss (DL). BCE is one of the most commonly used loss functions for machine learning in two-class classification tasks. For current work, the mask of each location is a zero or one matrix, which makes the task similar to a pixel-wise binary class classification. Therefore, BCE would be an appropriate loss function to use. The formula for BCE is shown below, considering the ground truth mask *gt* and the model predicted mask *pr*:

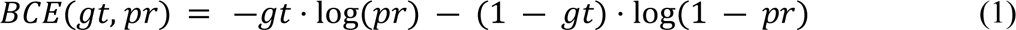

Another widely used loss function in segmentation tasks is the numeric sum of binary cross entropy and IoU score (Jaccard loss).

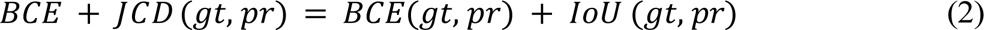

The Dice coefficient (DC) is a commonly used metric to calculate similarities between images. The Dice coefficient is defined similarly as IoU:

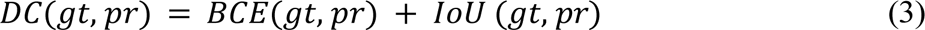

We split our dataset into training and validation sets. The training set contains 50% of the radiographs, and the validation contains the other 50%. For optimizing the hyperparameters, we used five-fold cross-validation with all possible combinations of hyper-parameters, including the optimizer, loss function, batch size, and learning rate. The list of tuning spaces for each hyper-parameter is shown in **Table 1**. To search through the proposed space of hyper-parameters, we used a Bayesian optimization process through the *scikit-optimize* package. We first defined an objective function that took instances of hyper-parameters, trained the model, and returned the cross-validation scores (CV scores). We then passed the scores to the optimization function of the package. The optimization process assumed the objective function results to follow a multivariate Gaussian distribution. It would take all observed scores until the current iteration, calculate a posterior distribution, and sample the next set of hyper-parameters instances out of the posterior distribution. The best combination of hyper-parameters is chosen for the final model training.

**Table 1.**
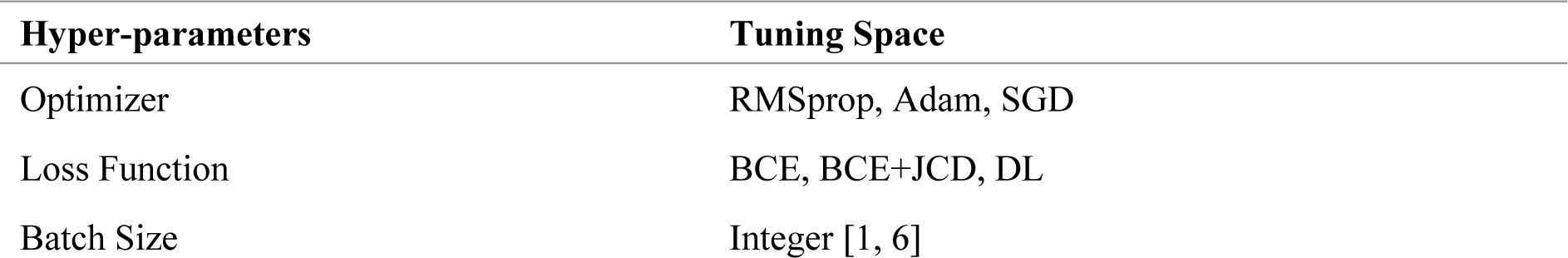
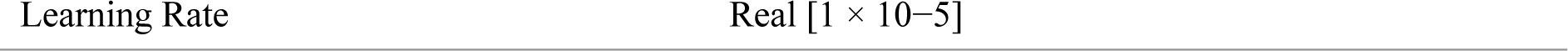
Search space for hyper-parameters. The categorical and integer variables (optimizer, loss function, and batch size) were initialized to have a uniform prior probability; the learning rate was initialized to have uniform prior distribution in log space (log-uniform). *RMSprop: root mean squared propagation; Adam: adaptive Moment Estimation; SGD: stochastic gradient descent; BCE: binary cross entropy; BCE+JCD: binary cross entropy with Jaccard loss; DL: Dice loss*.

### 2.5. Model Evaluation and Interpretability

We used intersection over union (IoU) and the Dice coefficient (i.e., F-score, Dice score) to evaluate. IoU, also known as Jaccard loss, is a commonly used metric in image segmentation tasks. Consider the ground truth mask *gt* and the model predicted mask *pr*:

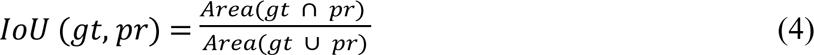

We assume that both masks are image matrices of 0’s and 1’s. Therefore, the area of the mask would be a count of 1’s in the corresponding pixel matrix. A high IoU score indicates that more pixels are predicted correctly (more true positives) while fewer pixels are missed (less false negatives and false positives). F-score represents a weighted average between precision and recall. Specifically,

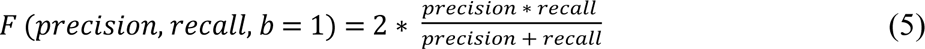

After obtaining the optimized hyperparameters, we fitted models using the original images and two distinct pre-processing techniques (i.e. CLAHE, and ps-KDE) onto the five anatomic structures (i.e., heart, left lung, right lung, left clavicle, right clavicle) with the corresponding best-performing hyperparameters for that task, for a total of 15 models. Our predictive models were then trained with 50 epochs, with 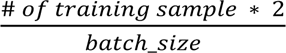 samples in each step. The validation loss is defined as the loss value when validated on the validating images during the last epoch. Within each dataset, we performed independent samples *t-test* assuming no equal variance using *R* (version 4.2.3). The significance level (*p*=0.01) was not corrected for multiple comparisons as none of the comparisons was tested more than once.

To understand our models’ classification, we randomly chose subjects and obtained the probability of each pixel being classified into the organ or clavicle. A heatmap was produced based on the probabilities using *Matplotlib*. In addition, we overlapped the model’s prediction with the original chest x-ray image to evaluate whether the segmentation has clinical merits.

## 3. Result

### 3.1. Cross-validation Results

The cross-validation results are shown in **Table 2**. The best loss function for all five locations was BCE+JCD, which considers pixel-wise information and intersection maximization.

**Table 2.** Cross-validation results. The best combination for each location is shown in the table. Note that for batch size, the actual batch size used in cross-validating and training models was the above batch size x 5. This multiplier was a result of the data augmentation, as we are loading the original image and augmented image all at the same time.

| Region | Optimizer | Loss Function | Batch Size | Learning Rate | CV Score |
| --- | --- | --- | --- | --- | --- |
| Heart | SGD | BCE+JCD | 1 | 0.07879 | 0.63121 |
| Left Clavicle | SGD | BCE+JCD | 1 | 0.10000 | 0.33311 |
| Left Lung | SGD | BCE+JCD | 1 | 0.00358 | 0.65930 |
| Right Clavicle | RMSprop | BCE+JCD | 1 | 0.00035 | 0.37285 |
| Right Lung | SGD | BCE+JCD | 1 | 0.00139 | 0.73626 |

### 3.2. ResNetUnet Evaluation

Using the original images (i.e., without CLAHE or ps-KDE transformation), the mean IoU ranged from 0.703 (SD=0.12) in the left clavicle, to 0.925 (SD=0.07) in the heart; the F1-score ranged from 0.537 (SD=0.28) in left clavicle to 0.918(SD=0.13) in the heart. Across the five models trained with CLAHE transformation, the mean IoU ranged from 0.666 (SD=0.14) in the right clavicle, to 0.921 (SD=0.08) in the heart; the F1-score ranged from 0.440 (SD=0.33) in right clavicle to 0.911 (SD=0.14) in the heart. Lastly, using ps-KDE transformation, the five models achieved mean IoU ranging from 0.577 (SD=0.06) in the right clavicle to 0.927 (SD=0.05) in the heart; the F1-score ranged from 0.275 (SD=0.17) in right clavicles to 0.926 (SD=0.070) in the heart **(Table 3**; **Figure 5**). In all three datasets, the best-performing model differed significantly from the worst-performing model (*p*<2.2 × 10^−16^).

**Figure 5.**
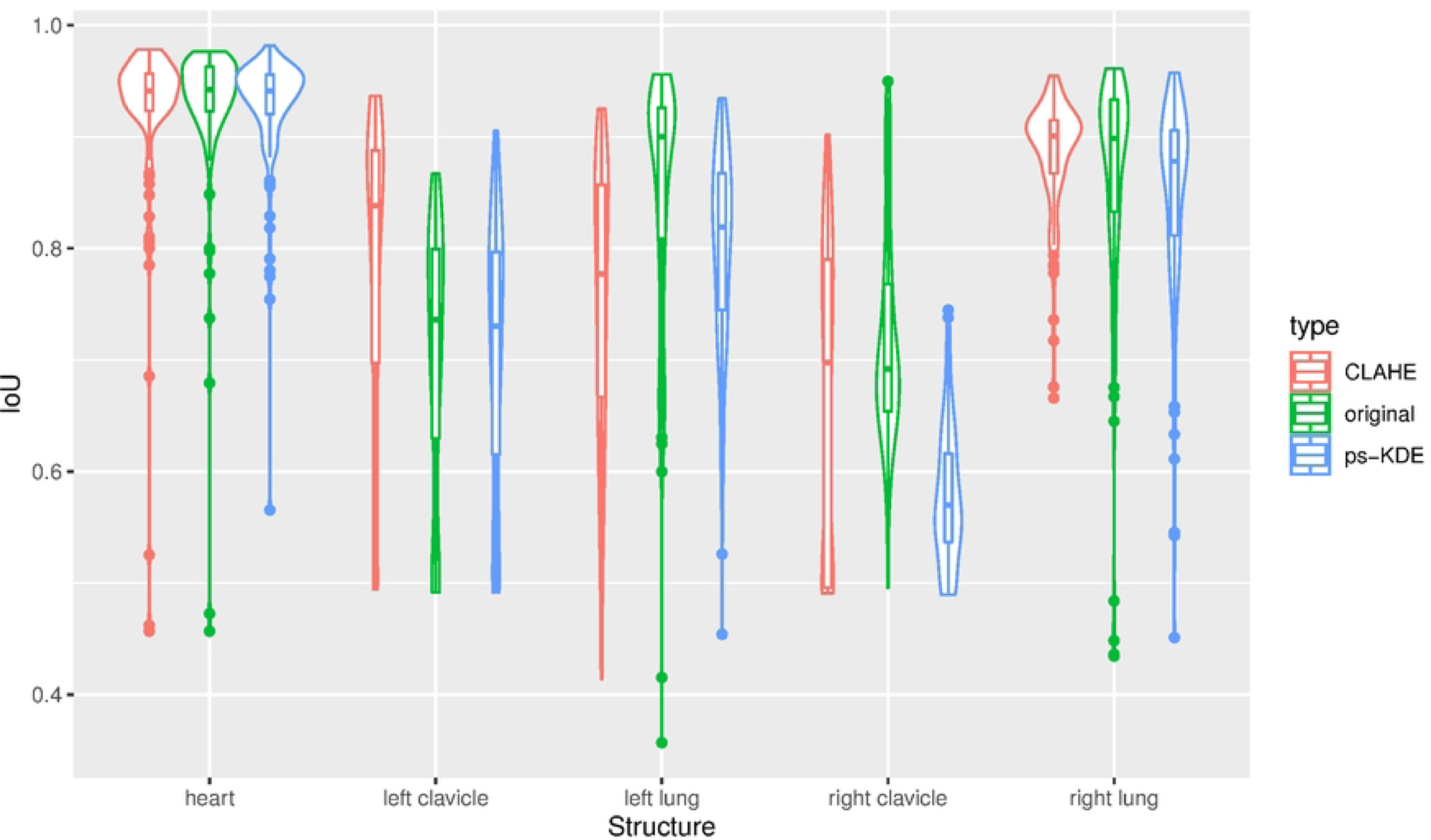
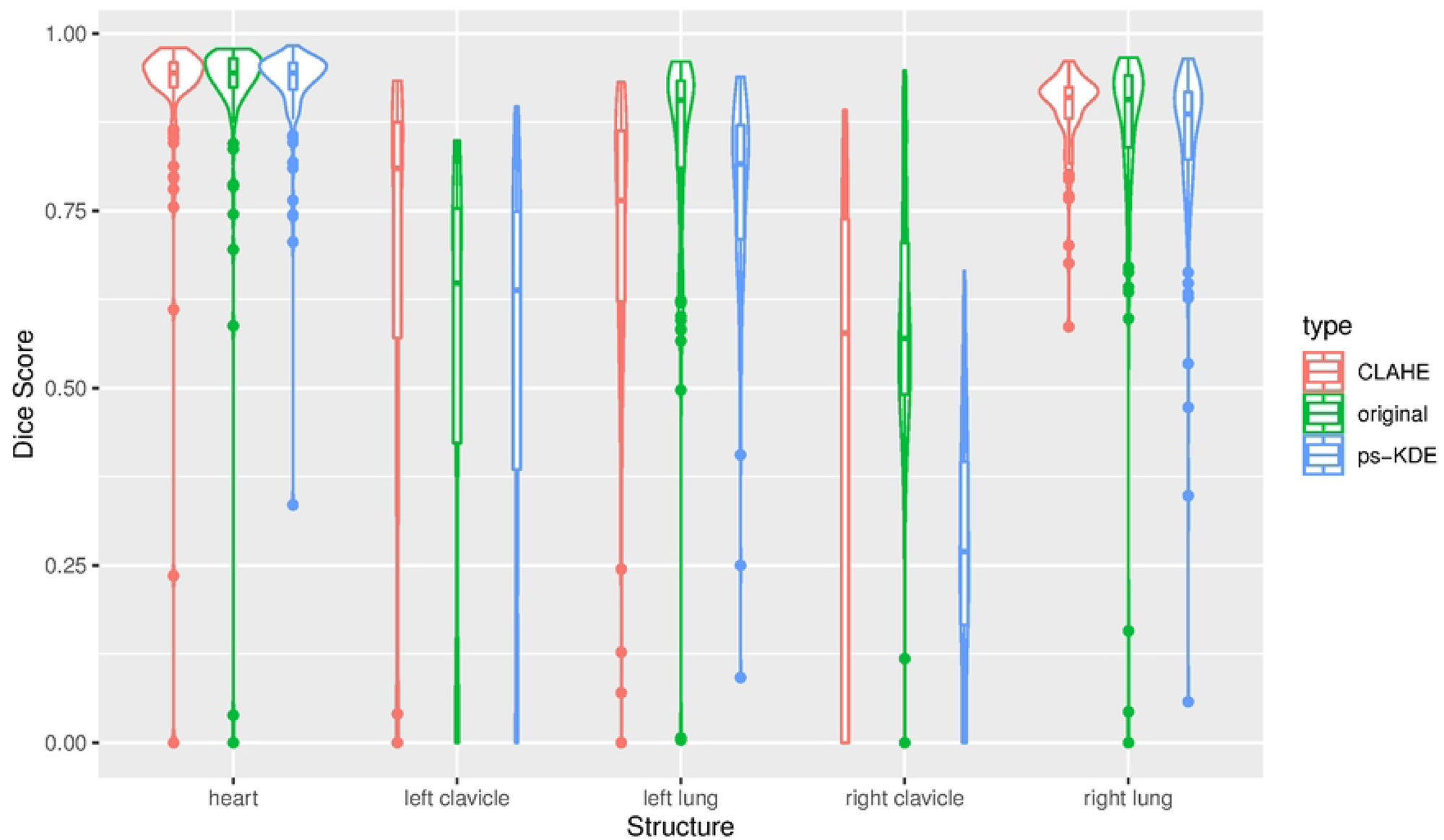
Model Performance Represented by Violin Plots for Each Anatomical Structure. a) IoU b) Dice score

**Table 3.** Model performance after applying preprocessing methods (CLAHE and ps-KDE) evaluated by IoU and Dice scores. IoU and Dice scores are shown as mean (SD). CLAHE: Contrast Limited Adaptive Histogram Equalization; Ps-KDE: Pixel-wise substitution by Kernel Density Estimation; IoU: Intersection over Union. *:p<0.01 in model performance when comparing between CLAHE and ps-KDE for each segmentation region pair.

|  | Region | IoU | Dice Score |
| --- | --- | --- | --- |
| Original | Heart | 0.925 (0.07) | 0.918 (0.13) |
|  | Left Clavicle | 0.703 (0.12) | 0.537 (0.28) |
|  | Left Lung | 0.848 (0.11) | 0.839 (0.16) |
|  | Right Clavicle | 0.716 (0.09) | 0.597 (0.17) |
|  | Right Lung | 0.863 (0.10) | 0.855 (0.17) |
| CLAHE | Heart | 0.921 (0.08) | 0.911 (0.14) |
|  | Left Clavicle | 0.778 (0.14) | 0.667 (0.30)* |
|  | Left Lung | 0.752 (0.12) | 0.717 (0.19)* |
|  | Right Clavicle | 0.666 (0.14) | 0.440 (0.33)* |
|  | Right Lung | 0.885 (0.05) | 0.894 (0.06)* |
| ps-KDE | Heart | 0.927 (0.05) | 0.926 (0.07) |
|  | Left Clavicle | 0.706 (0.12) | 0.545 (0.28)* |
|  | Left Lung | 0.799 (0.09) | 0.780 (0.13)* |
|  | Right Clavicle | 0.577 (0.06) | 0.275 (0.17)* |
|  | Right Lung | 0.848 (0.09) | 0.850 (0.12)* |

Among the Dice score and mean IoU metrics, there is no difference in the ranked order of model performances. Therefore, we will present Dice score only as the five models achieved a lower performance compared to that measured by mean IoU. This is to give a conservative estimate of the effectiveness of ps-KDE. We observed significant differences in model performance between the regions classified using CLAHE and ps-KDE. Specifically, in the left lung region, CLAHE had a Dice score of 0.717 (SD=0.19), and ps-KDE had a Dice score of 0.780 (SD=0.13), *p*=0.0026 (**Table 3**). We observed no differences between the two datasets in heart segmentation. CLAHE transformation achieved a significant result than ps-KDE in the left clavicle, right clavicle, and right lung.

Examples of model predictions with both processing techniques (CLAHE and ps-KDE) are shown in **Figure 6**. The probability heatmaps showed a decrease in confidence around the edges of the segmentation object. This is more prevalent in the heart and the left clavicle model. Visually, the overlap of the predicted segmentation from ps-KDE and the original x-ray pinpoints the regions that radiologists typically focus on. The partial misclassification in the right lung from the CLAHE technique is discussed in later sections.

**Figure 6.**
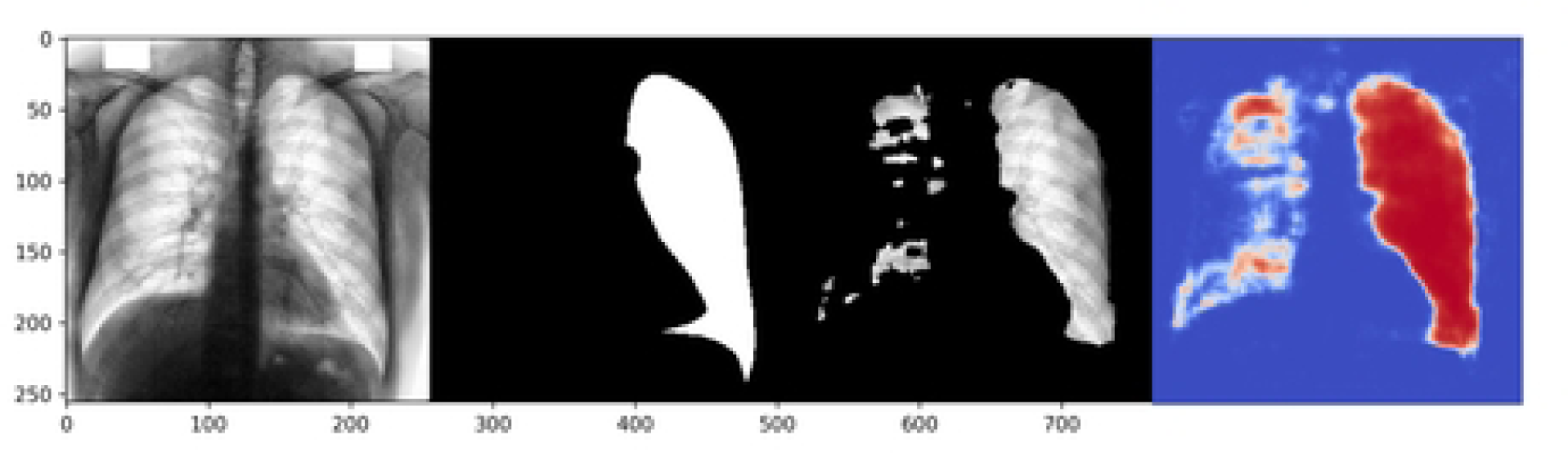
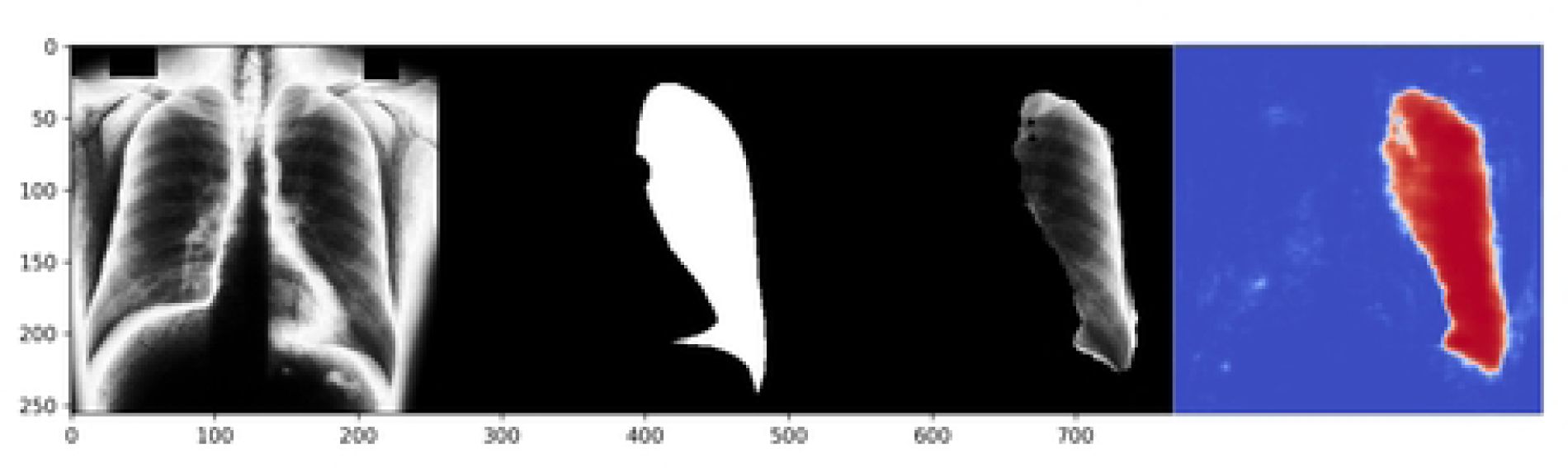
Prediction of a randomly selected subject. From left to right, the input of the model, the ground truth, the predicted segmentation overlap with the original x-ray, and the heatmap of the predicted probability. A) CLAHE processed, B) ps-KDE processed.

## 4. Discussion

In this study, we proposed a novel method, ps-KDE, to substitute the pixel value based on a normalized histogram distribution. Our investigation focused on evaluating the performance of the *ResNetUnet* architecture in the context of segmentation tasks, specifically applied to 247 chest X-rays with PA projection. We assessed each model’s segmentation capabilities across five distinct anatomic structures, considering the impact of preprocessing techniques such as ps-KDE and CLAHE.

In the original, CLAHE, and ps-KDE processed models, there exists a tremendous gap between the best and worst Dice scores. These fluctuations should be concerning as test images came from the same dataset. This suggests that although *U-Net* is supposedly designed for end-to-end biomedical image segmentation with very few samples, this algorithm with validation on electronic microscopy image stacks may not generalize to radiographs(10). We found that, in general, the model predicting lung regions and heart has the highest Dice scores, whereas in the clavicle regions, the Dice Score may drop below 0.6. The higher performance in large regions suggests the model could recognize larger patterns but fell short of smaller ones within the X-Ray.

We observed that models preprocessed with CLAHE have higher IoU and Dice scores (**Figure 5 (a, b)**) in left clavicle regions compared to the original image models. The ps-KDE method, on the other hand, showed better performance in the left lung model than CLAHE. This means in future studies we could explore the combined use of both preprocessing techniques through a dynamic voting algorithm, harnessing the advantage of CLAHE in smaller regions and that of ps-KDE in larger regions. The novelty of ps-KDE method lies within utilizing histogram values not only to generate density estimations but also to execute substitutions. Therefore, such combination allows the pixel substitution to benefit from CLAHE which has a more uniform overall distribution. By enabling accurate and consistent identification of anatomical structures, our proposed technique stands to enhance the precision of subsequent disease detection algorithms.

The incorporation of heatmaps offers invaluable insights into areas of interest and uncertainty during the segmentation process. Notably, we observed a consistent decrease in probability around object edges in the majority of images. This gradual phasing out of probability as the model progresses into negative pixels is ideal, as models exhibiting abrupt switches between high and low confidence levels may lack stability. The visualization of heatmaps also serves to pinpoint regions requiring further investigation. For instance, in CLAHE models, a few misclassifications of the right lung were observed when predicting left lung regions (**Figure 6**). This may be attributed to image augmentation techniques such as horizontal flips and rotation ranges applied before inputting the images. We hypothesize that, given the small size of our dataset, the spatial distribution of the ground truth significantly influences segmentation outcomes. This suggests that ps-KDE may exhibit greater robustness against substantial image augmentation and small datasets. Future studies could investigate the potential of applying transfer learning to *ResNetUnet* to mitigate the unintended impacts of augmentation(26, 27).

It’s worth noting that the predicted lungs still adhere to the clinical expectation that the left lung is narrow and long. Even in the case of the misclassified instance, we can observe that the model still accurately outlines the shape and conforms to the expected characteristics of the right lung. Heatmaps have the potential to empower clinicians by visually assessing segmentation accuracy and quality, facilitating interpretation and informed clinical decision-making.

## 5. Limitation

Our current dataset contains exclusively PNG images, whereas clinical practices heavily rely on the DICOM format for medical image analysis. While PNG is suitable for research and imaging information in DICOM can be easily converted to PNG format, it lacks the crucial metadata and standardized structure that DICOM would offer. This disconnection hinders the model’s direct applicability in clinical settings where DICOM’s comprehensive patient information and imaging details are essential.

To mitigate this limitation, the model needs further adaptation for DICOM data format. This involves adjusting the data processing pipeline to handle DICOM images and accounting for metadata intricacies. The model’s effectiveness must be re-validated using DICOM data to ensure its reliability in clinical workflows. Addressing this constraint is vital to bridge the gap between research-oriented PNG images and the practical demands of medical professionals who predominantly rely on DICOM for accurate diagnosis and treatment.

We also recognize that the size of our dataset is small for a deep learning algorithm. We also only trained *ResNetUnet* on 50 epochs because of computing resource constraints. Higher performance may be achieved in larger epochs. In addition, the smoothed histogram takes account of only the pixel distribution for this dataset. An additional limitation of our study is the absence of external validation for our models. From a dataset perspective, it remains uncertain how effectively the smoothed histograms can extend to external radiographs. Moreover, there is a potential for another enhanced *U-Net* architecture(28) to provide further validation regarding the applicability of the ps-KDE technique across various model architectures.

## 6. Conclusion

In conclusion, we significantly improved semantic segmentation of the left lung in chest radiographs using ps-KDE. The introduction of the ps-KDE preprocessing technique contributes to the available image contrasting methods for segmentation but should be treated with caution and further validations.

## Data Availability

All radiographs are held in a public repository from the Japanese Society of Radiological Technology (JSRT) database (http://db.jsrt.or.jp/eng.php)

## 7. Acknowledgments

We thank Kun-Hsing Yu, MD, Ph.D. for assistance with the segmentation algorithm, and Andrew Beam, Ph.D. for comments that greatly improved the experiment design. This work is inspired by the course BMI 707: Deep Learning for Biomedical Data. https://hms-dbmi.github.io/BMI_707/.

## 8. Declaration of Competing Interest

We, the authors of this paper, declare that we have no conflicts of interest related to the research presented in this manuscript. There are no financial interests or personal relationships that could be perceived as influencing the content within this work.

## Notes

### Competing Interest Statement

The authors have declared no competing interest.

### Funding Statement

The author(s) received no specific funding for this work.

## References

1. Chan HP, Samala RK, Hadjiiski LM, Zhou C. Deep Learning in Medical Image Analysis. Adv Exp Med Biol. 2020;1213:3–21.

2. Currie G, Hawk KE, Rohren E, Vial A, Klein R. Machine Learning and Deep Learning in Medical Imaging: Intelligent Imaging. J Med Imaging Radiat Sci. 2019;50(4):477–87.

3. Chen X, Wang X, Zhang K, Fung KM, Thai TC, Moore K, et al. Recent advances and clinical applications of deep learning in medical image analysis. Med Image Anal. 2022;79:102444.

4. Al-Antari MA, Al-Masni MA, Choi MT, Han SM, Kim TS. A fully integrated computer-aided diagnosis system for digital X-ray mammograms via deep learning detection, segmentation, and classification. Int J Med Inform. 2018;117:44–54.

5. Balkenende L, Teuwen J, Mann RM. Application of Deep Learning in Breast Cancer Imaging. Semin Nucl Med. 2022;52(5):584–96.

6. Masood A, Sheng B, Li P, Hou X, Wei X, Qin J, et al. Computer-Assisted Decision Support System in Pulmonary Cancer detection and stage classification on CT images. J Biomed Inform. 2018;79:117–28.

7. Sahiner B, Pezeshk A, Hadjiiski LM, Wang X, Drukker K, Cha KH, et al. Deep learning in medical imaging and radiation therapy. Med Phys. 2019;46(1):e1–e36.

8. Li Z, Hou Z, Chen C, Hao Z, An Y, Liang S, et al. Automatic cardiothoracic ratio calculation with deep learning. IEEE Access. 2019;7:37749–56.

9. Long J, Shelhamer E, Darrell T, editors. Fully convolutional networks for semantic segmentation. Proceedings of the IEEE conference on computer vision and pattern recognition; 2015.

10. Ronneberger O, Fischer P, Brox T, editors. U-net: Convolutional networks for biomedical image segmentation. Medical Image Computing and Computer-Assisted Intervention–MICCAI 2015: 18th International Conference, Munich, Germany, October 5-9, 2015, Proceedings, Part III 18; 2015: Springer.

11. Krithika Alias AnbuDevi M, Suganthi K. Review of Semantic Segmentation of Medical Images Using Modified Architectures of UNET. Diagnostics (Basel). 2022;12(12).

12. Wang S, Yang DM, Rong R, Zhan X, Xiao G. Pathology Image Analysis Using Segmentation Deep Learning Algorithms. Am J Pathol. 2019;189(9):1686–98.

13. Lee J, Pant SR, Lee H-S. An adaptive histogram equalization based local technique for contrast preserving image enhancement. International Journal of Fuzzy Logic and Intelligent Systems. 2015;15(1):35–44.

14. Li Y, Wang W, Yu D, editors. Application of adaptive histogram equalization to x-ray chest images. Second International Conference on Optoelectronic Science and Engineering’94; 1994: Spie.

15. Zimmerman JB, Pizer SM, Staab EV, Perry JR, McCartney W, Brenton BC. An evaluation of the effectiveness of adaptive histogram equalization for contrast enhancement. IEEE Transactions on Medical Imaging. 1988;7(4):304–12.

16. Pizer SM, Johnston RE, Ericksen JP, Yankaskas BC, Muller KE. Contrast-limited adaptive histogram equalization: speed and effectiveness. 1990. p. 337–45.

17. Alwakid G, Gouda W, Humayun M. Deep Learning-Based Prediction of Diabetic Retinopathy Using CLAHE and ESRGAN for Enhancement. Healthcare (Basel). 2023;11(6).

18. Yoshimi Y, Mine Y, Ito S, Takeda S, Okazaki S, Nakamoto T, et al. Image preprocessing with contrast-limited adaptive histogram equalization improves the segmentation performance of deep learning for the articular disk of the temporomandibular joint on magnetic resonance images. Oral Surg Oral Med Oral Pathol Oral Radiol. 2023.

19. Tjoa EA, Suparta IPYN, Magdalena R, CP NK, editors. The use of CLAHE for improving an accuracy of CNN architecture for detecting pneumonia. SHS Web of Conferences; 2022: EDP Sciences.

20. Shiraishi J, Katsuragawa S, Ikezoe J, Matsumoto T, Kobayashi T, Komatsu K, et al. Development of a digital image database for chest radiographs with and without a lung nodule: receiver operating characteristic analysis of radiologists’ detection of pulmonary nodules. AJR Am J Roentgenol. 2000;174(1):71–4.

21. van Ginneken B, Stegmann MB, Loog M. Segmentation of anatomical structures in chest radiographs using supervised methods: a comparative study on a public database. Med Image Anal. 2006;10(1):19–40.

22. Abadi M, Agarwal A, Barham P, Brevdo E, Chen Z, Citro C, et al. Tensorflow: Large-scale machine learning on heterogeneous distributed systems. arXiv preprint arXiv:160304467. 2016.

23. Pizer SM, Amburn EP, Austin JD, Cromartie R, Geselowitz A, Greer T, et al. Adaptive histogram equalization and its variations. Computer vision, graphics, and image processing. 1987;39(3):355–68.

24. He K, Zhang X, Ren S, Sun J, editors. Deep residual learning for image recognition. Proceedings of the IEEE conference on computer vision and pattern recognition; 2016.

25. Liu S, Deng W, editors. Very deep convolutional neural network based image classification using small training sample size. 2015 3rd IAPR Asian conference on pattern recognition (ACPR); 2015: IEEE.

26. Wang S, Rong R, Gu Z, Fujimoto J, Zhan X, Xie Y, et al. Unsupervised domain adaptation for nuclei segmentation: Adapting from hematoxylin & eosin stained slides to immunohistochemistry stained slides using a curriculum approach. Computer Methods and Programs in Biomedicine. 2023;241:107768.

27. Ganin Y, Lempitsky V, editors. Unsupervised domain adaptation by backpropagation. International conference on machine learning; 2015: PMLR.

28. Liu W, Luo J, Yang Y, Wang W, Deng J, Yu L. Automatic lung segmentation in chest X-ray images using improved U-Net. Sci Rep. 2022;12(1):8649.

